# METHYLATION BIOMARKERS ASSOCIATED WITH DRUG-RESISTANT EPILEPSY

**DOI:** 10.1101/2022.03.14.22271975

**Authors:** Patricia Sánchez Jiménez, Marcos Elizalde-Horcada, Ancor Sanz-García, Inmaculada Granero-Cremades, María De Toledo, Paloma Pulido, Marta Navas, Ana Beatriz Gago-Veiga, Lola Alonso-Guirado, María Concepción Alonso-Cerezo, Desirée Nava-Cedeño, Francisco Abad-Santos, Cristina Virginia Torres-Díaz, María Carmen Ovejero-Benito

## Abstract

Epilepsy is a disabling neurological disease that affects 2% of the population. Drug-resistant epilepsy (DRE) affects 25-30% of epilepsy patients. Understanding its underlying mechanisms is key to adequately manage this condition. To analyze the main epigenetic marks of DRE an epigenome-wide association study was carried out including samples from different regions of DRE patients’ brain and peripheral blood. An Illumina Infinium MethylationEPIC BeadChip array including cortex, hippocampus, amygdala, and peripheral blood from DRE subjected to neurosurgical resection of the epileptogenic zone was used. Overall, 32, 59, 3210, and 6 differentially methylated probes (DMPs) associated with DRE were found in the hippocampus, amygdala, cortex, and peripheral blood, respectively. These DMPs harbored 19, 28, 1574, and 7 genes, respectively, which play different roles in processes such as neurotrophic or calcium signaling. Three of the top DMPs observed in cortex were validated with methylation specific qPCR. Moreover, 163 DMPs associated with neurosurgery response at 6 months were found in the hippocampus. Genes located on these DMPs were involved in diverse processes such as synaptic signaling and central nervous system development. Besides 3 DMPs in blood samples were associated with response to neurosurgery at 12 months. In conclusion, the present study reports genome-wide DNA methylation changes across different regions of the DRE brain. These changes could be useful for further studies to disentangle the bases of DRE to search for therapeutic alternatives for this disease. Furthermore, they could also help identify patients likely to respond to neurosurgery.

## INTRODUCTION

New anticonvulsant drugs (ACs) have been approved for epilepsy in previous years. Nevertheless, 25-30% of epileptic patients are drug-resistant and this percentage is difficult to reduce^1^. Thus, drug-resistant epilepsy (DRE) is common, costly, and disabling^1^ and poses a therapeutic challenge, decreasing patients’ quality of life and increasing mortality^2^. Temporal lobe epilepsy (TLE) is often associated with DRE and can progress with hippocampal sclerosis that involves massive neuronal loss, and mossy fiber sprouting^3^. Neurosurgical resection of the epileptogenic zone is an effective way to achieve seizure freedom. Indeed, two-thirds of the patients achieve Engel I/II response after neurosurgery^4,5^. Conceivably early detection of DRE could further improve the success rate.

DRE underlying mechanisms are not fully characterized. Studying the basis of DRE is key to predict poor responses to ACs and to search for new therapeutic alternatives. Diverse mechanisms have been proposed to explain the processes occurring in the epileptogenic zone: genetic mechanisms, disease-related mechanisms and drug-related mechanisms and which may be interlinked^6^. Nevertheless, none of them could fully explain the disease^6^. As certain patients could respond to ACs and later become resistant to treatment^7^, epigenomic mechanisms may be involved in DRE^8^. In patients with TLE, drug-resistance appears, on average, nine years after starting pharmacological treatment^9^. This late onset may be explained by the involvement of environmental factors and epigenetic mechanisms in the development of drug resistance.

Epigenetic factors are stable but reversible modifications that may alter gene expression without affecting DNA sequence. Different epigenetic mechanisms are involved in epilepsy: DNA methylation, histone modifications, chromatin remodeling, and non-coding RNAs^10,11^. DNA methylation is the most stable epigenetic mark and is the most studied in DRE^10–23^. Hence, previous studies have found methylation differences associated with epilepsy in blood^13^ and brain samples^16,20^. Moreover, previous research found epigenetic biomarkers of focal cortical dysplasia^23^.

This study analyzed DNA methylation differences associated with DRE in different brain regions and peripheral blood samples to understand distinct processes underlying DRE and search for putative biomarkers of DRE. Besides, biomarkers of neurosurgery outcome were also analyzed.

## MATERIALS AND METHODS

### Study population and ethics statement

The protocol and the Informed Consent Form were approved by the Independent Clinical Research Ethics Committee of Hospital Universitario de La Princesa. The study was conducted following the Revised Declaration of Helsinki and STROBE guidelines. Blood and brain samples were obtained from DRE patients subjected to neurosurgical resection of the epileptogenic zone who signed the informed consent. The epileptogenic zone and the cortical surrounding area were collected from each patient. When the resection involved the amygdala, this tissue was also analyzed. Brain samples were frozen in dry ice immediately after their surgical extraction. An extensive analysis of the patients’ clinical records was carried out. The recruitment period started in September 2018 and lasted two years.

Outcomes of neurosurgical resection were evaluated using Engel classification^24^ at 6, 12, and 24 months after surgery. Patients were classified as responders (R) if they achieved Engel I and II and non-responders (NR) if they had Engel III and IV. Postmortem tissue of healthy subjects served as control brain samples. Cortex, hippocampus and amygdala samples were provided by the biobank “Biobanco en Red de la Región de Murcia”, BIOBANC-MUR. Blood control samples were obtained from “Biobanco del Hospital Universitario de La Princesa”. Control samples were processed following standard operating procedures with appropriate approval of the Ethics and Scientific Committees.

### Sample processing

DNA was extracted from peripheral blood samples using the MagNa Pure LC 2.0 System (Roche Applied Science, Germany). DNA and RNA were extracted from brain tissue samples (cortex, hippocampus, or amygdala) using the Allprep DNA/RNA Mini Kit according to the manufacturer’s protocol (Qiagen, Germany). DNA was quantified with Qubit dsDNA BR (Broad-Range) Assay Kit (Thermo Fisher Scientific, USA) for Invitrogen™ Qubit™ 4 Fluorometer (Thermo Fisher Scientific, USA).

### Genome-wide DNA methylation analysis with high-density arrays

The EZ DNA Methylation Kit (Zymo Research, USA) was applied for bisulfite conversion of 600ng of genomic DNA of the 95 DNA samples obtained from DRE patients. Genome-wide DNA methylation assay was carried out with the Infinium MethylationEPIC BeadChip Kit (Illumina Inc, California, USA) according to the manufacturer’s instructions^25^. Bisulfite-treated genomic DNA was whole-genome amplified, hybridized to Infinium MethylationEPIC BeadChips (Illumina, USA), and scanned using the Illumina iScan platform. The intensity of the images was extracted with the GenomeStudio Methylation Software Module (v1.9.0, Illumina, USA). Bisulfite and genotype experiments were performed by CEGEN (Centro Nacional de Genotipado, Madrid, Spain). Samples were randomized to avoid the batch effect. DNA extracted from some brain tissue samples was not enough to carry out this methylation analysis. Therefore, certain results could not be obtained for some samples

### Methylation preprocessing and analysis

The IDAT files generated from the Illumina MethylationEPIC BeadChip array were analyzed with the statistical software R (version 4.0.0) following a pipeline built on the minfi packages^26^, as described in Figure 1 and Supplementary Text. The final dataset comprised 772,002 CpG sites with good hybridization quality in 95 samples. Downstream analysis is detailed in Figure 1 and Supplementary Text.

**Figure 1:**
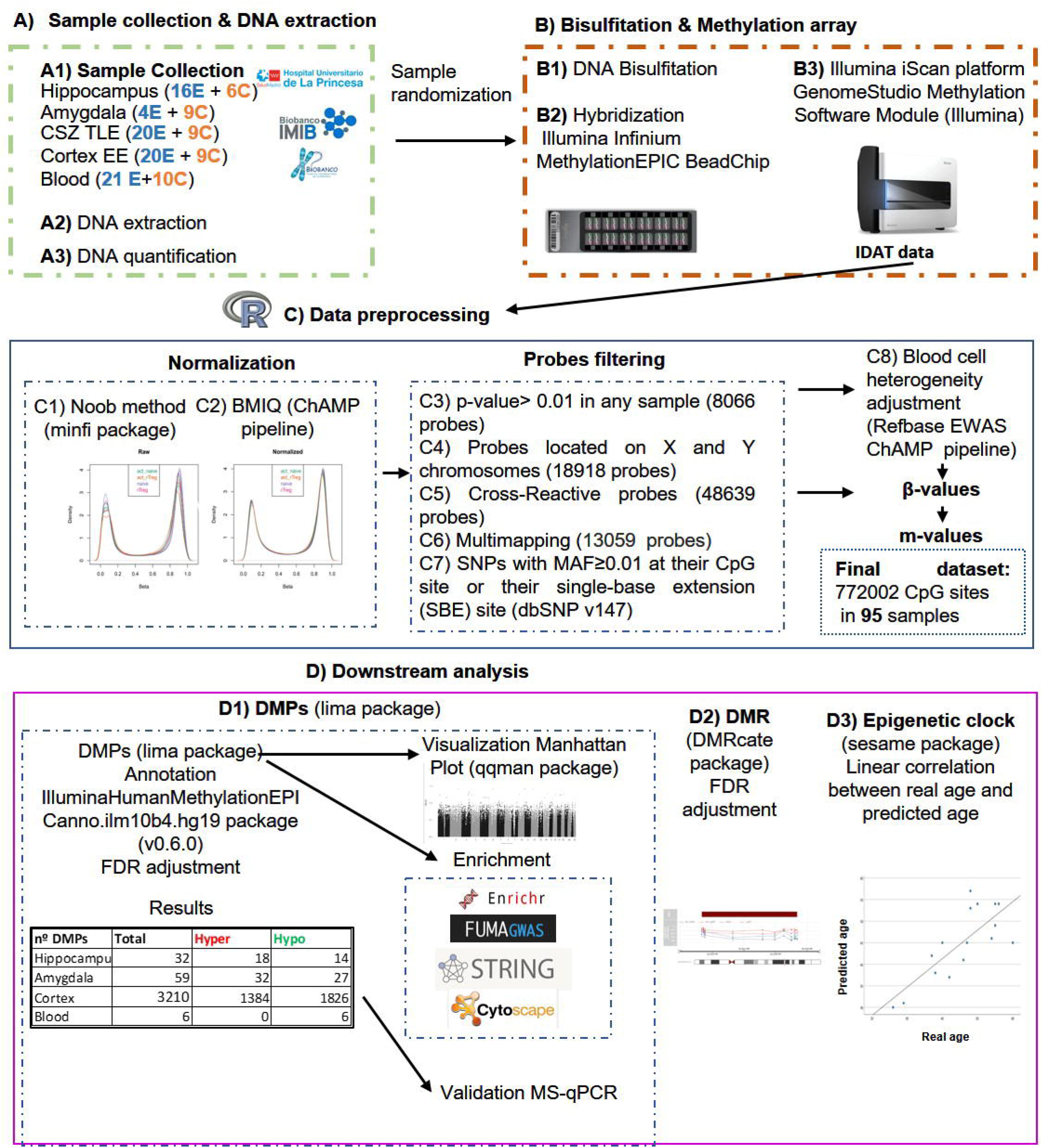
Workflow of the experiments and analysis carried out. A) Sample collection & DNA extraction. The number of drug resistant epilepsy patients (E) or controls (C) samples analyzed appear in brackets. B) Bisulfitation and methylation array. C) Data preprocessing. The numbers of probes that were discarded at each stage of the probes filtering are shown in brackets. D) Downstream analysis. The table that appears in the D1 section summarizes the DMPs observed in the different tissues analyzed. Abbreviations: C: controls, CSZ: cortical surrounding zone; DMPs: differentially methylated probes, DMR: differentially methylated regions; E: epilepsy; EE: extratemporal epilepsies; FDR: false discovery rate; Hyper: hypermethylated in patients with respect to controls; Hypo: Hypomethylated in patients with respect to controls; MS-qPCR: Methylation-specific qPCR NR, non-responders; R, responders; TLE: temporal lobe epilepsy.

### Methylation-specific qPCR (MS-qPCR)

DNA bisulfite conversion and qPCR conditions can be found in Supplementary Text. MS-qPCRs were analyzed as described^27^. Differentially methylation rate was calculated between patients and controls fold-change.

## RESULTS

### Study population

Twenty-three DRE patients subjected to neurosurgical resection of the epileptogenic zone were recruited (Table 1). Four of them presented extratemporal epilepsy and 19 TLE whose epileptogenic zone was located on the cortex. Most of the patients (95%) achieved Engel I/II 6 months after neurosurgery and thus were considered responders (Table 1). Ten controls were recruited for blood samples and another ten for brain postmortem tissue (Table 1).

**Table 1:** Summary of the clinical and demographic characteristics of the study population and the controls.

|  |  | DRE | Controls (brain) | Controls (blood) |
| --- | --- | --- | --- | --- |
| Women (%) |  | 9 (39%) | 3 (30%) | 3 (30%) |
| Age (years) |  | 46 ± 9 | 60 ± 12 | 57 ± 9 |
| Age at onset (years) |  | 18 ± 14 | NA | NA |
| Febrile seizures in childhood |  | 2 (9%) | NA | NA |
| Number of seizures/month |  | 7.3 ± 8.5 | NA | NA |
| Aura |  | 10 (43%) | NA | NA |
| n° anticonvulsant drugs |  | 5.4 ± 3.3 | NA | NA |
| Left dominant hand |  | 3 (13%) | NA | NA |
| Stress |  | 5 (22%) | NA | NA |
| Traumatism |  | 2 (9%) | NA | NA |
| Sensory, gait, or developmental disturbances |  | 4 (17%) | NA | NA |
| Neurosurgical hemisphere | Left | 13 (57%) | NA | NA |
|  | Right | 10 (42%) | NA | NA |
| Seizure etiology | Structural | 5 (22%) | NA | NA |
|  | Infectious | 2 (9%) | NA | NA |
|  | Unknown | 16 (69%) | NA | NA |
| Ever smoker |  | 11 (48%) | NA | NA |
| Alcohol |  | 8 (34%) | NA | NA |
| Engel (1 month) | R | 20 (87%) | NA | NA |
|  | NR | 3 (13%) | NA | NA |
| Severe adverse effects (1m) |  | 6 (26%) | NA | NA |
| Engel (6 months) | R | 21 (95%) | NA | NA |
|  | NR | 2 (5%) | NA | NA |
| Severe adverse effects (6m) |  | 4 (17%) | NA | NA |
| Engel (12 months) | R | 21 (95%) | NA | NA |
|  | NR | 2 (5%) | NA | NA |
| Severe adverse effects (12m) |  | 3 (13%) | NA | NA |
| Engel (24 months) |  | 11 (85%) | NA | NA |
|  |  | 2 (5%) | NA | NA |
| Severe adverse effects (24m) |  | 4 (17%) | NA | NA |

Data are shown as mean ± SD or number and percentage. As blood samples were not available from the subjects who donated their brains, control blood samples were taken from a different group of control subjects. Accordingly, two different cohorts of controls were used: control (blood) and control (brains)
Abbreviations: DRE, drug resistant epilepsy; NA, not applicable; NR, non-responders; R, responders.

### DMPs associated with epilepsy

#### Hippocampus

Hippocampal samples containing the epileptogenic zone of DRE patients (N=16) were compared in the methylation analysis with control hippocampus (N=8) to study changes occurring in the epileptogenic zone of DRE patients. A total of 32 significant DMPs harboring 19 different genes were identified (Figure 2A, Table 2, Supplementary Table 1A). The main functions of these genes included sympathetic nervous system development, and central nervous system development, among others (Supplementary Figure 1 and Supplementary Table 2A).

**Table 2:**
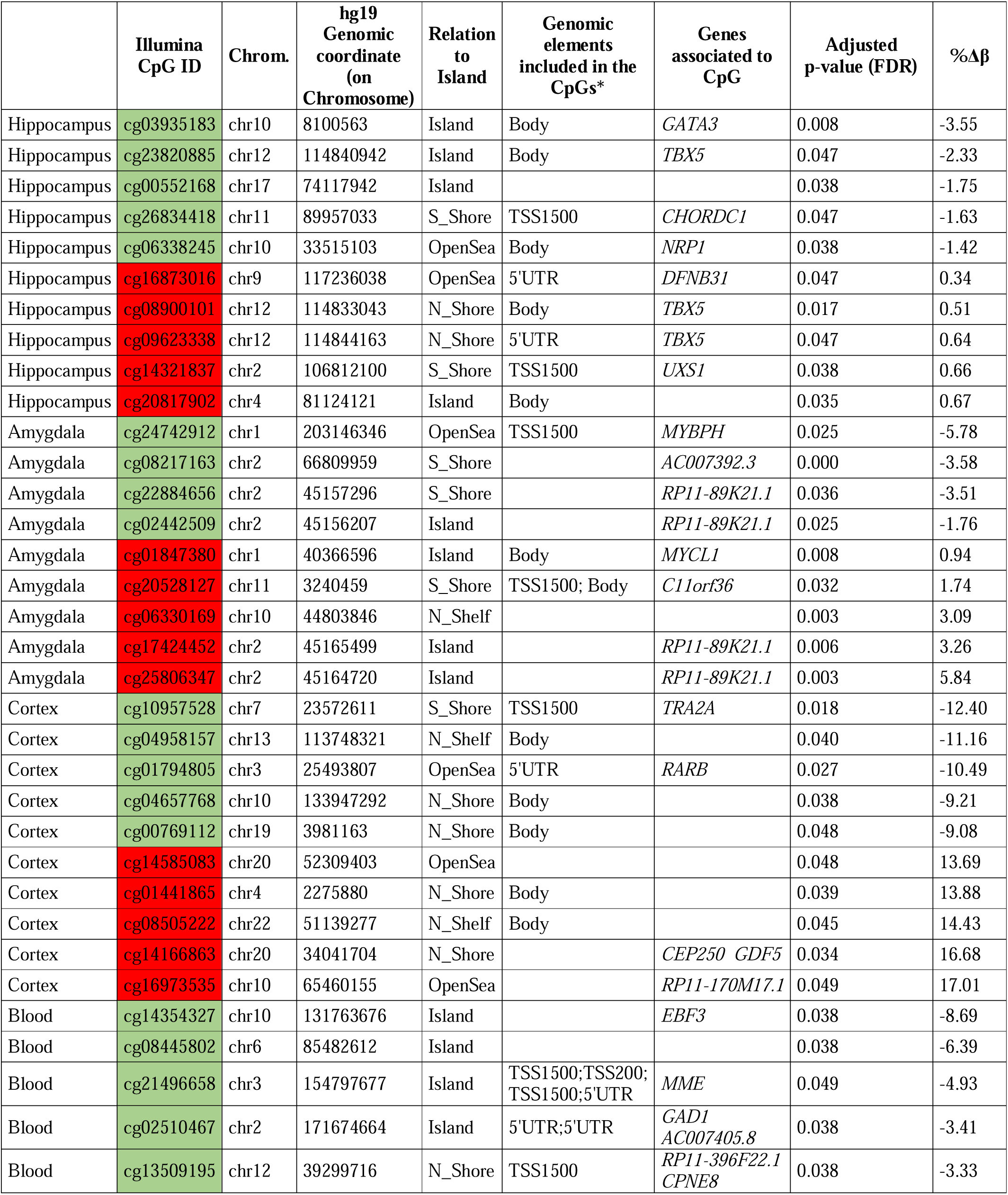
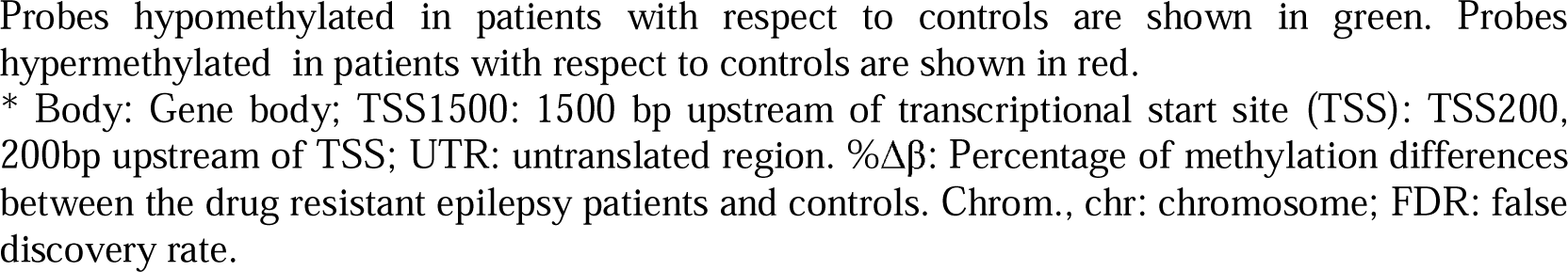
Summary of the top 5 hypomethylated and hypermethylated genes in different brain areas.

| | Illumina CpG ID | Chrom. | hg19 Genomic coordinate (on Chromosome) | Relation to Island | Genomic elements included in the CpGs* | Genes associated to CpG | Adjusted p-value (FDR) | %$\Delta\beta$ |
| --- | --- | --- | --- | --- | --- | --- | --- | --- |
| Hippocampus | cg03935183 | chr10 | 8100563 | Island | Body | *GATA3* | 0.008 | -3.55 |
| Hippocampus | cg23820885 | chr12 | 114840942 | Island | Body | *TBX5* | 0.047 | -2.33 |
| Hippocampus | cg00552168 | chr17 | 74117942 | Island |  |  | 0.038 | -1.75 |
| Hippocampus | cg26834418 | chr11 | 89957033 | S_Shore | TSS1500 | *CHORDC1* | 0.047 | -1.63 |
| Hippocampus | cg06338245 | chr10 | 33515103 | OpenSea | Body | *NRP1* | 0.038 | -1.42 |
| Hippocampus | cg16873016 | chr9 | 117236038 | OpenSea | 5'UTR | *DFNB31* | 0.047 | 0.34 |
| Hippocampus | cg08900101 | chr12 | 114833043 | N_Shore | Body | *TBX5* | 0.017 | 0.51 |
| Hippocampus | cg09623338 | chr12 | 114844163 | N_Shore | 5'UTR | *TBX5* | 0.047 | 0.64 |
| Hippocampus | cg14321837 | chr2 | 106812100 | S_Shore | TSS1500 | *UXS1* | 0.038 | 0.66 |
| Hippocampus | cg20817902 | chr4 | 81124121 | Island | Body |  | 0.035 | 0.67 |
| Amygdala | cg24742912 | chr1 | 203146346 | OpenSea | TSS1500 | *MYBPH* | 0.025 | -5.78 |
| Amygdala | cg08217163 | chr2 | 66809959 | S_Shore |  | *AC007392.3* | 0.000 | -3.58 |
| Amygdala | cg22884656 | chr2 | 45157296 | S_Shore |  | *RP11-89K21.1* | 0.036 | -3.51 |
| Amygdala | cg02442509 | chr2 | 45156207 | Island |  | *RP11-89K21.1* | 0.025 | -1.76 |
| Amygdala | cg01847380 | chr1 | 40366596 | Island | Body | *MYCL1* | 0.008 | 0.94 |
| Amygdala | cg20528127 | chr11 | 3240459 | S_Shore | TSS1500; Body | *C11orf36* | 0.032 | 1.74 |
| Amygdala | cg06330169 | chr10 | 44803846 | N_Shelf |  |  | 0.003 | 3.09 |
| Amygdala | cg17424452 | chr2 | 45165499 | Island |  | *RP11-89K21.1* | 0.006 | 3.26 |
| Amygdala | cg25806347 | chr2 | 45164720 | Island |  | *RP11-89K21.1* | 0.003 | 5.84 |
| Cortex | cg10957528 | chr7 | 23572611 | S_Shore | TSS1500 | *TRA2A* | 0.018 | -12.40 |
| Cortex | cg04958157 | chr13 | 113748321 | N_Shelf | Body |  | 0.040 | -11.16 |
| Cortex | cg01794805 | chr3 | 25493807 | OpenSea | 5'UTR | *RARB* | 0.027 | -10.49 |
| Cortex | cg04657768 | chr10 | 133947292 | N_Shore | Body |  | 0.038 | -9.21 |
| Cortex | cg00769112 | chr19 | 3981163 | N_Shore | Body |  | 0.048 | -9.08 |
| Cortex | cg14585083 | chr20 | 52309403 | OpenSea |  |  | 0.048 | 13.69 |
| Cortex | cg01441865 | chr4 | 2275880 | N_Shore | Body |  | 0.039 | 13.88 |
| Cortex | cg08505222 | chr22 | 51139277 | N_Shelf | Body |  | 0.045 | 14.43 |
| Cortex | cg14166863 | chr20 | 34041704 | N_Shore |  | *CEP250 GDF5* | 0.034 | 16.68 |
| Cortex | cg16973535 | chr10 | 65460155 | OpenSea |  | *RP11-170M17.1* | 0.049 | 17.01 |
| Blood | cg14354327 | chr10 | 131763676 | Island |  | *EBF3* | 0.038 | -8.69 |
| Blood | cg08445802 | chr6 | 85482612 | Island |  |  | 0.038 | -6.39 |
| Blood | cg21496658 | chr3 | 154797677 | Island | TSS1500;TSS200; TSS1500;5'UTR | *MME* | 0.049 | -4.93 |
| Blood | cg02510467 | chr2 | 171674664 | Island | 5'UTR;5'UTR | *GAD1 AC007405.8* | 0.038 | -3.41 |
| Blood | cg13509195 | chr12 | 39299716 | N_Shore | TSS1500 | *RP11-396F22.1 CPNE8* | 0.038 | -3.33 |

Probes hypomethylated in patients with respect to controls are shown in green. Probes hypermethylated in patients with respect to controls are shown in red.
\* Body: Gene body; TSS1500: 1500 bp upstream of transcriptional start site (TSS); TSS200, 200bp upstream of TSS; UTR: untranslated region. % $\Delta\beta$ : Percentage of methylation differences between the drug resistant epilepsy patients and controls. Chrom., chr: chromosome; FDR: false discovery rate.

**Figure 2.**
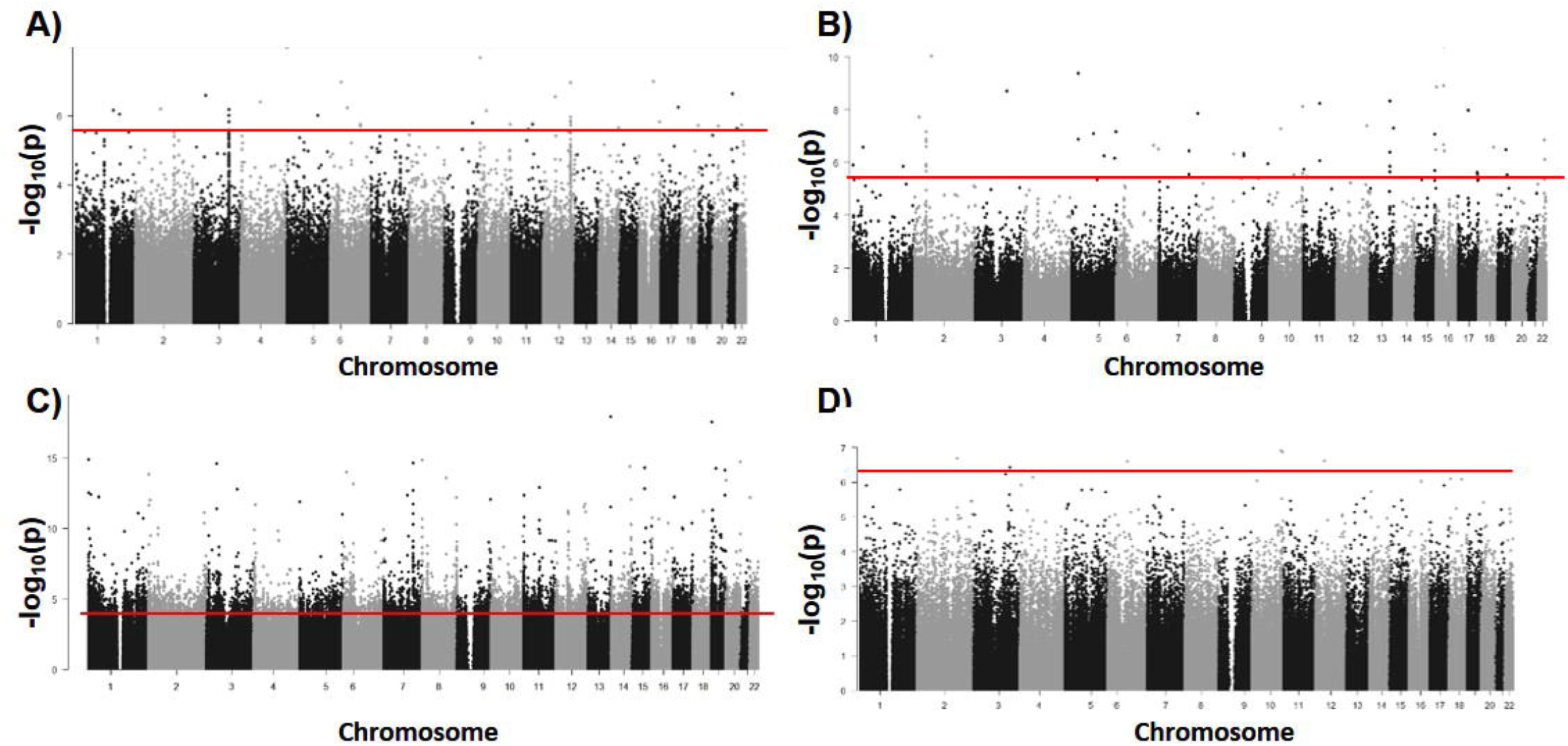
Manhattan plot of DMPs associated with DRE. Genomic coordinates are displayed along the X-axis, with the negative logarithm of the association p-value for each CpG site displayed along the Y-axis. Points below the horizontal line are not statistically significant. A) Hippocampus, B) Amygdala, C) Surrounding cortex to the epileptogenic zone, D) Peripheral blood after adjusting by the different cell types.

#### Amygdala

In several DRE patients, the epileptogenic zone covered both the hippocampus and the amygdala. In this case, DMPs were also analyzed in amygdala samples from these patients (N=4) and controls (N=9), finding 59 DMPs associated with DRE in this tissue (Figure 2B, Table 2, Supplementary Table 1B). These DMPs were annotated to 28 genes, which were involved in diverse processes such as neuroinflammatory cascades or neurotrophic factor signaling (Supplementary Table 2B).

#### Cortex

To access the hippocampal epileptogenic zone, the anterolateral temporal lobe was partially resected. To explore potential specific epigenetic modifications in that surrounding cortex to the epileptogenic zone (SCEZ), we compared samples from DRE patients (N=16) and controls (N=9). We found 3210 significant DMPs associated with DRE (Figure 2C, Table 2, Supplementary Table 1C), which harbor 1574 different genes involved in diverse biologic functions (Supplementary Table 2C and Supplementary Figure 2A and B). Proteins encoded by those genes were grouped in clusters of genes related with specific functions (Supplementary Table 3). Supplementary Figure 3 shows the 4 largest clusters. Cluster 1 includes 81 proteins involved in functions such as actin filament organization, and filamentous actin (Supplementary Figure 3A). Cluster 2 is composed by 23 proteins voltage-gated channel, and Transient receptor potential channels (Supplementary Figure 3B). Cluster 3 comprises 20 proteins related to the spliceosome (Supplementary Figure 3C). Cluster 4 contains 16 proteins involved in DNA repair (Supplementary Figure 3D).

**Table 3:** Summary of hypomethylated and hypermethylated genes validation by MS-qPCR in the surrounding cortex to the epileptogenic zone.

| Gene |  | Drug resistant epilepsy Patients | Healthy Controls | Methylation Rate |
| --- | --- | --- | --- | --- |
| ***SHANK3*** | $\Delta\Delta CT$ Average | $-33.35 \pm 2.10$ | $-32.92 \pm 1.76$ | 1.34<br>(1.06 – 1.71) |
| | Fold Change | $1.09 \cdot 10^{10}$<br>($2.54 \cdot 10^{09} - 4.69 \cdot 10^{10}$) | $8.12 \cdot 10^{09}$<br>($2.39 \cdot 10^{09} - 2.75 \cdot 10^{10}$) | |
| ***SBF1*** | $\Delta\Delta CT$ Average | $-38.50 \pm 1.61$ | $-33.77 \pm 1.63$ | 26.71<br>(27.19 – 26.23) |
| | Fold Change | $2.90 \cdot 10^{11}$<br>($1.28 \cdot 10^{11} - 1.19 \cdot 10^{12}$) | $1.46 \cdot 10^{10}$<br>($4.71 \cdot 10^{09} - 4.54 \cdot 10^{10}$) | |
| ***MCF2L*** | $\Delta\Delta CT$ Average | $-30.01 \pm 1.31$ | $-31.09 \pm 2.13$ | 0.47<br>(0.84 – 0.27) |
| | Fold Change | $1.08 \cdot 10^{09}$<br>($4.37 \cdot 10^{08} - 2.68 \cdot 10^{09}$) | $2.29 \cdot 10^{09}$<br>($5.22 \cdot 10^{08} - 1.01 \cdot 10^{10}$) | |

Data are shown as mean ± SD or fold-change and its range for each gene. The fold-change range was calculated as $2^{-(\Delta\Delta CT - SD)}$ and $2^{-(\Delta\Delta CT + SD)}$ . Methylation rates were calculated as a ratio between patients and controls fold-change. The results were expressed as the fold change and 95% confidence interval.
As there was a fold-change range, methylation rate had a confidence range. SD: standard deviation.

No DMPs were found between the cortical epileptogenic zone (N=4) and controls (N=9) in extratemporal epilepsy. Additionally, no DMPs were observed between the cortical region of patients whose epileptogenic zone was in the cortex (N=4) and the SCEZ (N=16).

#### Blood

Peripheral blood samples were obtained from DRE patients and controls to detect putative surrogate biomarkers of DRE that could help predict drug-resistant epilepsy in a non-invasive manner. Blood samples from DRE (N=21) were compared with controls (N=10) finding 19,780 CpGs (7,615 hypermethylated, 12,165 hypomethylated) (data not shown). Nevertheless, as methylation is cell-type specific, the effect of cell type specific differences was corrected considering differences in patients’ blood cell populations. Afterward, only 6 significant DMPs remained, all of them hypomethylated in patients compared to controls (Figure 2D, Table 2, Supplementary Table 1D).

#### Validation by MS-qPCR

Three of the top DMPs located in SCEZ that presented an increment of methylation beta values between patients and controls higher than 10% and that were located on the gene body were selected for validation. We confirmed with an alternative technique (MS-qPCR) that *SHANK3* and *ZFYVE28* were hypermethylated in patients with respect to controls (Table 3). Moreover, we observed that *MCF2L* presented a ratio lower than 1 and thus, is hypomethylated in patients with respect to controls (Table 3). These results are consistent with those obtained in the methylation array (Supplementary Table 1C).

#### DMRs associated with epilepsy

Apart from performing a genome-wide analysis, DMRs analysis was carried out to determine whether several proximal CpGs were concordantly differentially methylated. Three DMRs harboring 3 genes were found in the hippocampus and 12 DMRs harboring 11 genes in the amygdala (Supplementary Table 4A-B).

Moreover, 531 significant DMRs associated with DRE harboring 500 genes were found in the SCEZ (Supplementary Table 4C). They play functions such as regulation of Rho activity, NGF, tight junctions, and PI3K/AKT activation (Supplementary Table 4D).

Further details on the results of the DMPs associated with the surgical outcome and the epigenetic clock calculations, can be found in Supplementary Text, Supplementary Figures 4 and 5, Supplementary Table 5.

## DISCUSSION

In the present study we analyzed differential methylation associated with DRE. Unlike previous studies^10–23^, we analyzed different tissues simultaneously, searched for markers of response to surgery, validated part of the results with an alternative technique and studied the epigenetic clock.

DMPs associated with DRE were analyzed in different regions of the central nervous system. A total of 32 significant DMPs associated with DRE harboring 19 different genes were found in the hippocampus. According to GO Biological Process 2019, they were involved in processes such as autonomic nervous system development, sympathetic nervous system development, and positive regulation of axon guidance (Supplementary Table 2, Supplementary Figure 1). A previous study conducted in the hippocampus (9 DRE and 5 controls) found 146 GO terms associated with development, neural remodeling, and neuron maturation^20^. The difference in the number of differentially methylated genes between this work^20^ an our study, may be partially explained by the different methodology used in both studies (Methylated DNA immunoprecipitation^20^ vs. methylation array) and the number of patients included in the studies (9 DRE and 5 controls^20^ vs. 16 DRE and 8 controls). Another study conducted in a rat model of TLE showed a predominant increase of methylation in the hippocampus^28^. These results are consistent with our findings, which showed hypermethylation in hippocampus and amygdala samples, whereas the cortex and blood showed hypomethylation.

Methylation studies following a candidate gene approach showed increased expression of DNA methyltransferases 1 and 3a (*DMT1* and *DMT3a*) was increased in human TLE^21^. According to previous studies^14,19,22^, we did not observe differences in methylation of Reelin, *EPHX1*, or *CPA6* in DRE. However, *ACAP3* was hypomethylated in the cortex, as described previously^15^.

Methylation analysis in the amygdala identified 59 DMPs harboring 28 genes. These genes are involved in processes such as PKC activation through G-Proteins, or signaling through both Trk and p75 (NTR) neurotrophin receptors, which are related to DRE^29^ Moreover, these genes also play roles in functions related to neuroinflammatory cascades such as IL1 mediated signaling or TNF receptor signaling, which are also associated with DRE^6^. To the best of our knowledge, this is the first study reporting methylation differences in the amygdala.

In the SCEZ, 1574 genes showed differences in methylation compared to control tissue. These genes are involved in processes previously associated with DRE, such as calcium signaling, neurotrophic signaling^29^, axon guidance, and long term potentiation (Supplementary Table 2C, Supplementary Figure 2A-B). The proteins encoded by these genes were grouped in clusters of genes related to specific functions. Interestingly, one of these cluster was associated with growth factors and kinases, and the second largest cluster consisted of Voltage-gated channels, the main targets of ACs (Supplementary Figure 3, Supplementary Table 3). This result suggests that these genes encoding ACs targets may be altered even in regions surrounding the epileptogenic zone. Moreover, several rare pathologies associated with these genes, such as encephalopathy recurrent of childhood or juvenile myoclonic epilepsy, are related to epilepsy (Supplementary Table 2C). A subset of 3 genes that showed DMPs in the SCEZ were validated with an alternative technique MS-qPCR. *SHANK3* and *SBF1* were hypermethylated and *MCF2L* was hypomethylated in DRE with respect to controls using both techniques thus confirming the reliability of DMPs obtained from the methylation array. These differentially methylated CpGs were classified into 546 DMRs (Supplementary Table 4C). Their associated functions are similar to those described by a previous study conducted in hippocampal samples^16^. (Supplementary Table 4D). Although the SCEZ has been often classified as “unaffected” tissue, the high implication of epigenetics reinforces the hypothesis of a neural network disease^3^, independently of the location of the epileptogenic zone. No DMPs were found between extratemporal epilepsies and the surrounding cortex to the epileptogenic zone. We also failed to find significant differences between the cortical epileptogenic zone and controls. The lack of differences may be caused by the small number of extratemporal samples (N=4) and their heterogeneity. Several DMPs associated to DRE were shared by different brain tissues. They are discussed in Supplementary Text.

Previous studies have also investigated DMPs in peripheral blood, finding 216 sites associated with epilepsy in white blood cells^13^. These DMPs harbor 130 genes: 47 of these DMPs match those observed in peripheral blood in the present study. However, after applying a correction for methylation differences due to variations in the proportion of blood cell subpopulations to avoid spurious associations, these coincidences disappeared leaving just 6 significant DMPs, all hypomethylated in patients compared to the controls.

Methylation changes reflecting epilepsy-associated changes observed in different regions of the brain analyzed (hippocampus, cortex, or amygdala) were not found in peripheral blood samples^30^. This may be explained by tissue and cell specificity of DNA methylation. Indeed, from the 32 DMPs significantly associated with epilepsy in hippocampal samples, only cg26834418 showed a strong correlation between blood and brain with BECon (r=0.56)^30^ and https://han-lab.org/methylation/default/imageCpG^31^ (p=0.004, r=0.61) (data not shown). This CpG could be proposed as potential non-invasive biomarker of DRE. However, it should be validated in a wider cohort of patients.

Epigenetic biomarkers of aging were measured in the different brain tissue samples from DRE patients to determine the effects of epilepsy on brain aging. The predicted epigenetic age was like the chronological or real age for the hippocampus and cortex, thus ruling out a potential acceleration of aging in the epileptogenic zone. Nevertheless, the predicted mean age of the amygdala was higher than its chronological age.

Neurosurgical resection of the epileptogenic zone is a successful therapeutic approach for treating DRE^4^. Despite the reduced number of patients who did not respond appropriately to the surgery (N=2), we found 163 DMPs harboring 114 genes associated with response to surgery (Supplementary Table 5A, Supplementary Figure 5B). They were involved in many processes related to epilepsy, such as nitric oxide signaling, axon guidance, developmental biology, excitatory chemical synaptic transmission, calcium ion transport into the cytosol, NMDA glutamate receptor activity, ionotropic glutamate receptor activity, ligand-gated calcium channel activity, glutamate receptor complex, and Partial epilepsy (Supplementary Table 5A). These processes are similar to those described previously^16^. Moreover, 3 DMPs were found between NR (N=2) and R (N=18) in the SCEZ (Supplementary Table 5C). These DMPs harbor the following genes: *C20orf27* and *SHTN1*. Shootin-1 (*SHTN1*) is involved in neuronal polarization and neurite outgrowth and is associated with developmental and Epileptic Encephalopathy 2. Furthermore, we detected two hypermethylated CpGs, (cg01006898 and cg13714644 [*DDX10*]) and one hypomethylated CpG (cg18484110) in NR compared to R in peripheral blood (Supplementary Table 5D). Although these genes might act as putative biomarkers of neurosurgical response, these results should be interpreted with caution due to the limited number of patients, and they must be confirmed in more extensive cohorts of DRE patients.

The study presents several limitations. First, differential methylation results may be affected by the reduced sample size of the groups of different tissues, such as the amygdala or the extratemporal epilepsy cortical samples, due to the low frequency of neurosurgical resections. Moreover, the quality of the brain samples in this study was high because they were frozen immediately after neurosurgical resection, and thus they were not subjected to fixation in formalin that could bias the results. Second, determination of the DMPs associated with the neurosurgical response may be affected by the higher number of patients who reached an Engel I or II score and the lower number of non-responders to neurosurgery. Therefore, these results should be interpreted with caution. Third, as blood samples from donors of postmortem control brain tissue were not available, a different cohort of control was included for blood samples. Four, the correction for cell type specific effects on methylation could not be applied to brain samples because the proportion of different cell subpopulations could not be properly assessed in brain samples. Since the biomarker available for this determination (NeuN) is expressed only in a subpopulation of cells in the hippocampus this adjustment could introduce a new bias in this study^33,34^. Finally, all patients were treated with ACs, which may influence DNA methylation status^17^. Taken all together, although our results provide interesting insights into the role of methylation epilepsy, further analysis should be performed to validate the results.

In conclusion, we have unveiled different epigenetic patterns associated with DRE in different regions of the brain. They provide interesting insights into epigenetic modifications potentially involved in DRE that could be candidate therapeutic targets. Besides, we have validated part of the results with an alternative technique. We also found potential biomarkers associated with surgery outcomes in peripheral blood. Further research is needed to confirm these results.

## Data Availability

All data produced in the present study are available upon reasonable request to the authors.

## Acknowledgments

We are particularly grateful for the generous contribution of the patients and the collaboration of Biobanco-HUP (Hospital Universitario de la Princesa) and Biobank Network of the Region of Murcia, BIOBANC-MUR, registered on the Registro Nacional de Biobancos with registration number B.0000859. BIOBANC-MUR is supported by the “Instituto de Salud Carlos III (proyecto PT20/00109), by “Instituto Murciano de Investigación Biosanitaria Virgen de la Arrixaca, IMIB” and by “Consejeria de Salud de la Comunidad Autónoma de la Región de Murcia”. We would like to thank Manuel Gómez Gutierrez for his help with the study and their valuable comments on this manuscript. The genotyping was performed at the Spanish National Cancer Research Centre, in the Human Genotyping lab, a member of CeGen, PRB3 and is supported by grant PT17/0019, of the PE I+D+i 2013-2016, funded by ISCIII and ERDF. We would like to thank Dr. Agustín Fernández-Fernández and Hortensia de la Fuente for their valuable advice.

## Data availability statement

All data produced in the present study are available upon reasonable request to the authors.

## FIGURES AND TABLES

**Supplementary Table 1**. Significant differentially methylated probes associated with drug resistant epilepsy in different tissues. A) Hippocampus, B) Amygdala, C) Surrounding cortex to the epileptogenic zone, D) Peripheral blood after adjusting by the different cell types. Probe location and the gene annotation were taken from Illumina reference files.

Body: Gene body; TSS1500: 1500 bp upstream of transcriptional start site (TSS): TSS200, 200bp upstream of TSS; UTR: untranslated region. %Δβ: Percentage of methylation differences between the drug resistant epilepsy patients and controls. chr: chromosome; FDR: false discovery rate.

Probes hypomethylated in patients with respect to controls are shown in green. Probes hypermethylated in patients with respect to controls are shown in red.

**Supplementary Table 2**. Enrichr of the significant differentially methylated probes associated with drug resistant epilepsy in different tissues. A) Hippocampus, B) Amygdala, C) Surrounding cortex to the epileptogenic zone.

**Supplementary Table 3**. Definition of the main clusters created by Cytoscape. Cluster 1 includes 81 proteins involved in functions such as actin filament organization, and filamentous actin. Cluster 2 is composed by 23 proteins Voltage-gated channel, and Transient receptor potential channels. Cluster 3 is formed by 20 proteins related to the spliceosome. Cluster 4 has 16 proteins involved in DNA repair. Proteins included in cluster 1 are shown in orange, cluster 2 in blue, cluster 3 in green and cluster 4 in yellow.

**Supplementary Table 4**. Significant differentially methylated regions found in the different tissues (A-C). A) Hippocampus, B) Amygdala, C) Surrounding cortex to the epileptogenic zone. D) Enrichr of the genes located on the differentially methylated regions located in the surrounding cortex to the epileptogenic zone.

Abbreviations: chr: chromosome; HMFDR: harmonic mean of the individual; meandiff: Mean differences in DNA methylation (%) between patients and controls are shown as a measurement of the effect size. Fisher <0.05 is considered significant.

**Supplementary Table 5A)** Significant differentially methylated probes associated with response to neurosurgery at 6 months in the hippocampus. 5B) Enrichr of differentially methylated probes associated with response to neurosurgery at 6 months in the hippocampus. 5C) Significant differentially methylated probes associated with response to neurosurgery at 6 months in the surrounding cortex to the epileptogenic zone. 5D) Significant differentially methylated probes associated with response to neurosurgery at 12 months in the peripheral blood after adjusting by the different cell types. Probe location and the gene annotation were taken from Illumina reference files.

Body: Gene body; TSS1500: 1500 bp upstream of transcriptional start site (TSS): TSS200, 200bp upstream of TSS; UTR: untranslated region. %Δβ: Percentage of methylation differences between the drug resistant epilepsy patients and controls. chr: chromosome; FDR: false discovery rate.

Probes hypomethylated in patients with respect to controls are shown in green. Probes hypermethylated in patients with respect to controls are shown in red.

**Supplementary Figure 1**. Enrichment analysis performed with FUMA GWAS analyzing functions of the genes involved in DMPs in the hippocampus according to the GO molecular function library. Red bars represent the proportion of overlapping genes in gene set. Blue bars show the enrichment p value, represented as the -logarithm of the FDR adjusted p value. Yellow squares show the genes involved in every enrichment term. Abbreviation: FDR: false discovery rate.

**Supplementary Figure 2**. Enrichment analysis performed with FUMA GWAS analyzing functions of the genes involved in DMPs in Cortical surrounding zone of patients compared with cortex of healthy controls. A) Biocarta. B) KEGG pathways. Red bars represent the proportion of overlapping genes in gene set. Blue bars show the enrichment p value, represented as the -logarithm of the FDR adjusted p value. Yellow squares show the genes involved in every enrichment term. Abbreviation: FDR: false discovery rate.

**Supplementary Figure 3**. Cytoscape networks of the top 4 largest clusters obtained with String. Cluster 1 includes 81 proteins involved in diverse functions such as actin filament organization, and filamentous actin. Cluster 2 is composed by 23 proteins related to voltage-gated channel, and Transient receptor potential channels. Cluster 3 is made up of 20 proteins related to the spliceosome. Cluster 4 has 16 proteins involved in DNA repair.

**Supplementary Figure 4**. Enrichment analysis performed with FUMA GWAS analyzing functions of the genes involved in DMPs associated with neurosurgery response at 6 months in the hippocampus according to KEGG pathways. Red bars represent the proportion of overlapping genes in gene set. Blue bars show the enrichment p value, represented as the - logarithm of the FDR adjusted p value. Yellow squares show the genes involved in every enrichment term. Abbreviation: FDR: false discovery rate.

**Supplementary Figure 5**. Correlation between drug resistant patients’ real age and predicted age by the epigenetic clock in the different tissues. A) Hippocampus, B) Amygdala, C) Surrounding cortex to the epileptogenic zone, D) Peripheral blood after adjusting by the different cell types. *p<0.05

