## Supplementary Text for "METHYLATION BIOMARKERS ASSOCIATED WITH DRUG-RESISTANT EPILEPSY"

#### Supplementary materials and methods

**Methylation preprocessing and analysis**

The IDAT files from the Illumina MethylationEPIC array were analyzed with the statistical software R (version 4.0.0) following a pipeline built on the minfi packages^1^. Initially, methylation data was normalized by the Noob method^2^. Extracted β-values were subsequently normalized using the BMIQ method^3^ implemented in ChAMP^4^ to correct for probe design. Quality control and correction of genetic variation was carried out by filtering the following probes: those with detection p-value>0.01 in any sample, sex chromosome probes, cross-reactive and multimapping probes^5,6^ and those with SNPs with MAF≥0.01 at their CpG site or their single-base extension (SBE) site (dbSNP v147). The final dataset is comprised of 772,002 CpG sites with good hybridization quality in 95 samples. M-values were obtained by the logit transformation of β-values. Methylation data sets were adjusted for blood cell heterogeneity using Refbase EWAS^7^.

Differentially methylated probes (DMPs) were calculated by the limma package^8^ which was used to compute a moderated t-test adjusted by the batch effect (the ratio of the m-value to its standard error). In order to determine the effect of the t-test on the magnitude of differential methylation, the effect size was calculated for each CpG site in EWAS performed with categorical variables (epilepsy vs controls or NR vs R). Raw p-values were corrected using the Benjamini–Hochberg multiple comparison procedure for false discovery rate (FDR) adj. p-value<0.05 was considered significant. Probes were annotated to genes and genomic locations by applying the IlluminaHumanMethylationEPICanno.ilm10b4.hg19 package (v0.6.0).

Manhattan plots were represented using the R packages qqman and manhattanlly.

**Differentially methylated regions (DMRs)**

DMRs were analyzed with the R package DMRcate^9^ as previously described^1^. DMRs with a Fisher p-value lower than 0.05 were considered significant.

**Enrichment analysis**

Enrichr (<https://amp.pharm.mssm.edu/Enrichr/>)^10,11^ and FUMA GWAS (<https://fuma.ctglab.nl/>) were applied to identify signaling pathways and biological functions associated with drug resistant epilepsy or the neurosurgical response. The main databases used were related to Pathways, Ontologies, and Diseases/Drugs. Only significant results with an adjusted p-value lower than 0.05 were considered.

String db (<https://string-db.org/>) and Cytoscape software were used to perform protein network analysis (Supplementary Figure 3).

**DNA Methylation Age Measures**

Predicted methylation age (DNAmAge) was calculated using the sesame R package^12^. Linear regression models were performed to analyze the association of chronological real age with the predicted epigenetic age.

**Methylation-specific qPCR (MS-qPCR).**

The EZ DNA Methylation Kit (Zymo Research, USA) was applied for bisulfite conversion of 1000ng of genomic DNA of two DNA cortex samples obtained from drug-resistant epilepsy patients and controls, respectively. Human Methylated & Non-methylated DNA Set (Zymo Research, USA) was employed for control of conversion reaction control and the Non-methylated DNA control of the set for standard curves of unmethylated primers.

| Gene | Primers | Sequence | Probes | Sequence (5'-3') |
| --- | --- | --- | --- | --- |
| MCF2L | FORWARD | GTATGGTTTTAGTTTTTCGGTT | METHYLATED | [6FAM]ACTAACAAAAAAATAACCCCCGAC[TAM] |
| *MCF2L* | REVERSE | CGCCCTTAACCTATAAAATACC | NO METHYLATED | [HEX]ACTAACAAAAAAATAACCCCCAAC[TAM] |
| *SBF1* | FORWARD | GTTTTTGTAGAGTTGTTTTGTTAG | METHYLATED | [6FAM]AATACGAACGAATAAAAACCACCC[TAM] |
| *SBF1* | REVERSE | AACCATAACTTACCTAACCTCCTA | NO METHYLATED | [HEX]AATACAAACAAATAAAAACCACCC[TAM] |
| *SHANK3* | FORWARD | AGTTATGGTGTGTGTGATTTGG | METHYLATED | [6FAM]TCTACCCCGAAACCTCTAACACT[TAM] |
| *SHANK3* | REVERSE | AAACACTATCACTACTCCCACTCC | NO METHYLATED | [HEX]TCTACCCCAAAACCTCTAACACT[TAM] |
| *ACTB* | FORWARD | TGGTGATGGAGGAGGTTTAGTAAGT |  | [6FAM]CACCACCCAACACACAATA[TAM] |
| *ACTB* | REVERSE | AACCAATAAAACCTACTCCTCCCTTAA |  |  |

As positive controls and standard curve of methylated primers, Bisulfite Converted Universal Methylated Human DNA Standard (Zymo Research, USA) was used. A subset of three genes, two hypermethylated (*SHANK3, SBF1*) and one hypomethylated (*MCF2L*) were selected from methylation array analysis. All presented an increment of methylation β values between DRE and controls higher than 10% in the surrounding cortex to the epileptogenic zone and the CpG was in the body of the gene. Actin B (*ACTB)* was used as a reference gene in the qPCR. A region without CpGs was selected for *ACTB* amplification. Thus, just one probe was designed for *ACTB*.

Reactions were performed with 10ng of genomic DNA in a final volume of 20μl, adjusted to 400nM of each primer (Custom DNA Oligos, Merck, Germany) and 200nM of each probe, methylated and unmethylated (Dual-Label Probe, Merck, Germany) and 1X Applied Biosystems ^TM^ TaqMan® Genotyping Master Mix (Thermo Fisher Scientific, USA). The qPCR cycling conditions were 60ºC for 30 s, 95ºC for 10 min, followed by 40 cycles for 15 s at 95ºC, 1 min at 60ºC with fluorescence measurement, 30 s at 60ºC. Real-time qPCR analysis was performed using QuantStudio^TM^ 12K Flex (Thermo Fisher Scientific, USA) and following the analysis described by^13^.

**Supplementary results**

**DMPs associated with response to neurosurgery**

The association of DMPs with response to neurosurgery was also analyzed in this study. (Table 1). A total of 163 DMPs were found between NR (N=2) and R (N=14) in the hippocampus (Supplementary Table 5A) after 6 months after neurosurgery. These CpGs harbor 114 genes that play different roles such as axon guidance and ionotropic glutamate receptor activity, among others (Supplementary Table 5B). In addition, 3 DMPs were found between NR (2) and R (18) in the cortex surrounding the epileptogenic zone (Supplementary Table 5C). Nevertheless, as the NR group included only 2 patients, these results should be interpreted with caution and should be validated in a different cohort of patients. Moreover, 3 DMPs associated with neurosurgery response at 12 months were also found in our study. These CpGs are different from those found associated with epilepsy in peripheral blood (Supplementary Table 5D).

**Epigenetic clock**

Epigenetic clock was calculated to analyze to determine if the epileptogenic zone is more/less aged than its chronological age would suggest. Epigenetic biomarkers of ageing were calculated in the diverse tissues obtained to determine the effects of these diseases on the ageing brain. Participants had a mean (SD) chronological age of 46±9 years. DRE patients’ predicted age was 50.3±6.7 (amygdala); 44.5±9.1 (Surrounding cortex to the epileptogenic zone) 45.1±9.77 (hippocampus) and 48.3±10.7 (blood). Chronological age was highly correlated with predicted age in hippocampus (ρ=0.779, p=0.000) and cortex (ρ=0.875, p=0.000) (Supplementary Figures 5A, B). Nevertheless, no significant correlation was found between predicted and chronological age in amygdala (ρ=0.875, p=0.125) and blood (ρ=0.125, p=0.590) (Supplementary Figures 5B, D).

#### Supplementary discussion

Although most DMPs were not simultaneously found in the cortex and hippocampus, one gene that plays a role in transcriptional regulation, *RP11-73O6.4* *LINC02157* (Long Intergenic Non-Protein Coding RNA 2157), showed DRE associated differential methylation in both brain regions. Furthermore, *RNF40* was also subjected to epigenetic regulation both in the cortex and amygdala. *RNF40* (Ring Finger Protein 40) encodes an E3 ubiquitin-protein ligase that facilitates the degradation of syntaxin 1 involved in the neurotransmitter release machinery. This result is in agreement with a study showing that ubiquitin ligases are upregulated in hippocampal sclerosis epilepsy samples^14^. Three more genes were hypomethylated both in the cortex and amygdala: *MPPED1, RP11-560I19.4* and *PRKCZ. MPPED1* (metallophosphoesterase domain-containing protein), which is highly expressed in the human fetal brain, may regulate the development or the function of cortical and hippocampal neurons through its metallophosphodiesterase activity^15^. *RP11-560I19.4* (FEZF1 antisense RNA 1), with yet an unknown function, seems to be related to schizophrenia. *PRKCZ* encodes a kinase involved in long-term potentiation. Consistently, a previous study reported a deletion in this gene in epilepsy patients^16^. Moreover, *LINC02157* was also hypermethylated in the cortex and amygdala. However, its function remains unknown and it has not been associated to DRE yet. Surprisingly, more DMPs were found the in cortical surrounding tissue than in the epileptogenic zone. These results may partially be explained by the cell death observed in the epileptogenic zone in TLE^17^ thereby suggesting that neurons subjected to methylation changes may die by apoptosis.
