## Supplementary figures and images for "METHYLATION BIOMARKERS ASSOCIATED WITH DRUG-RESISTANT EPILEPSY"

### Supplementary Figure 1

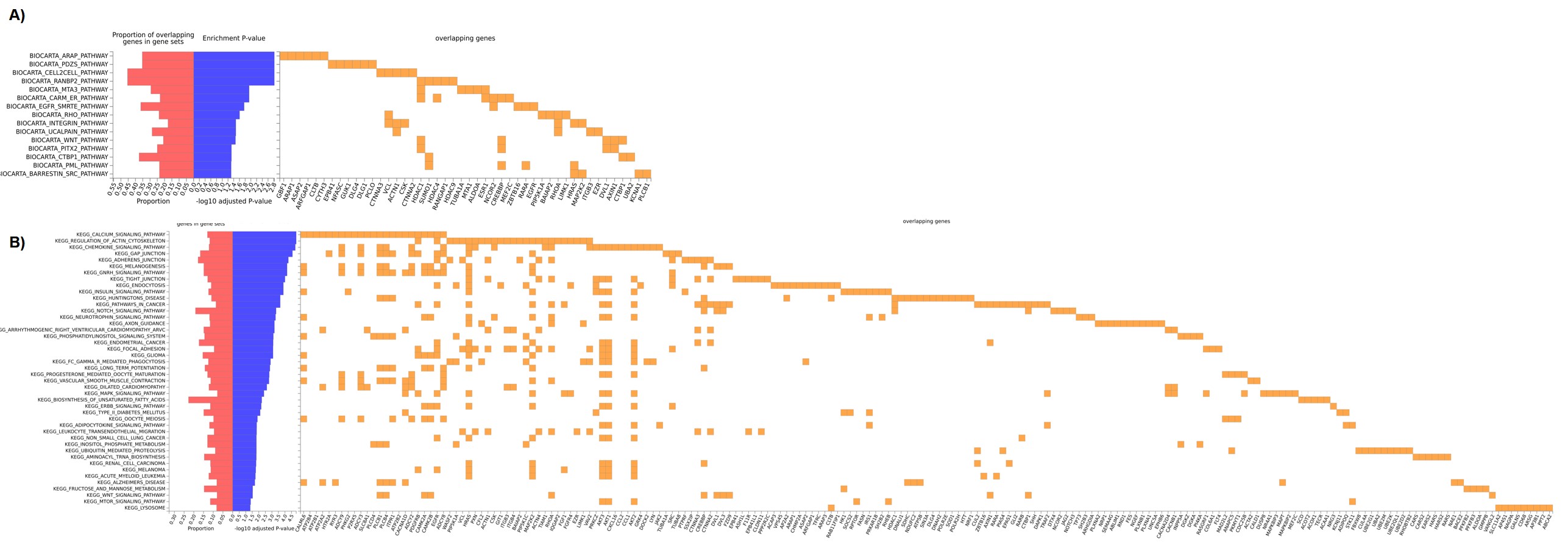

### Supplementary Figure 2

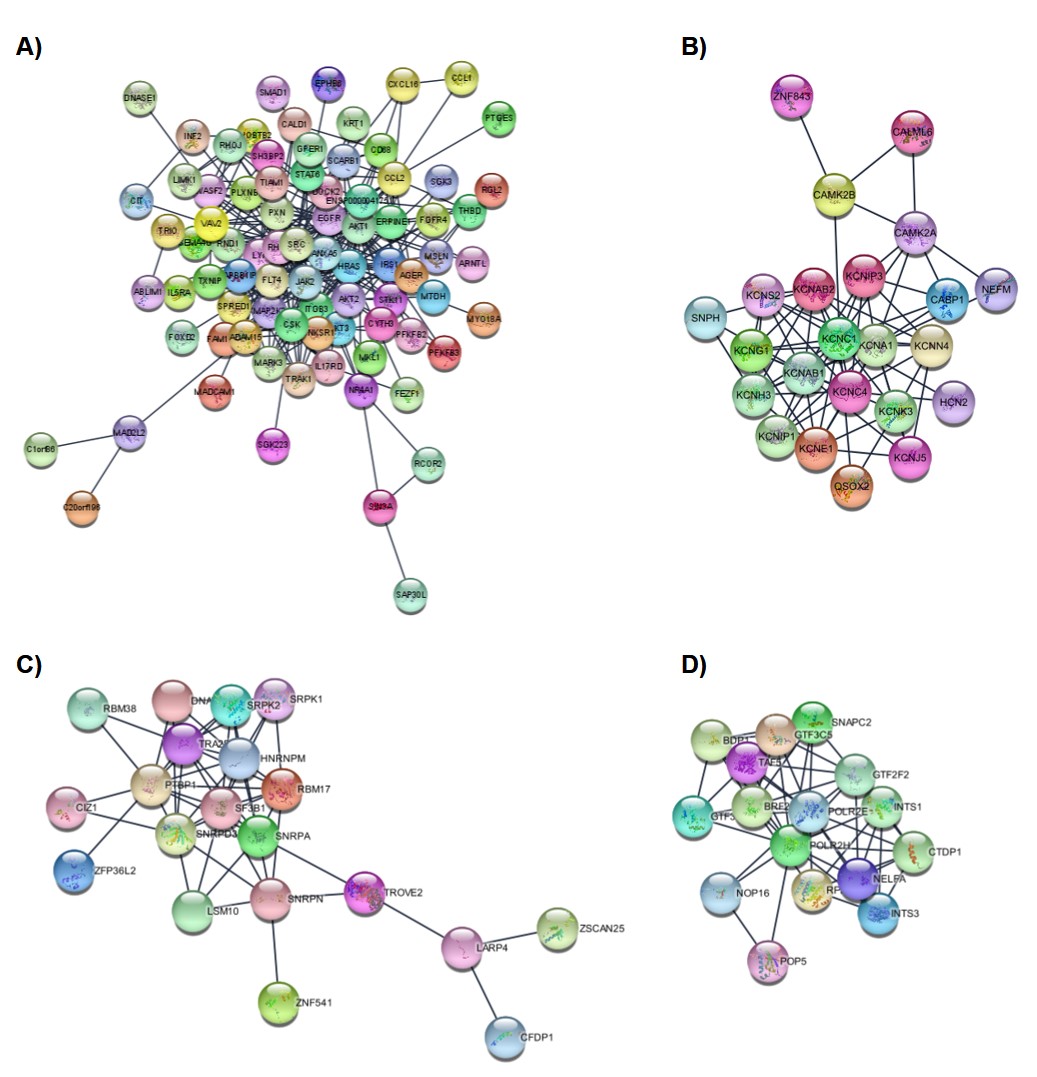

### Supplementary Figure 3

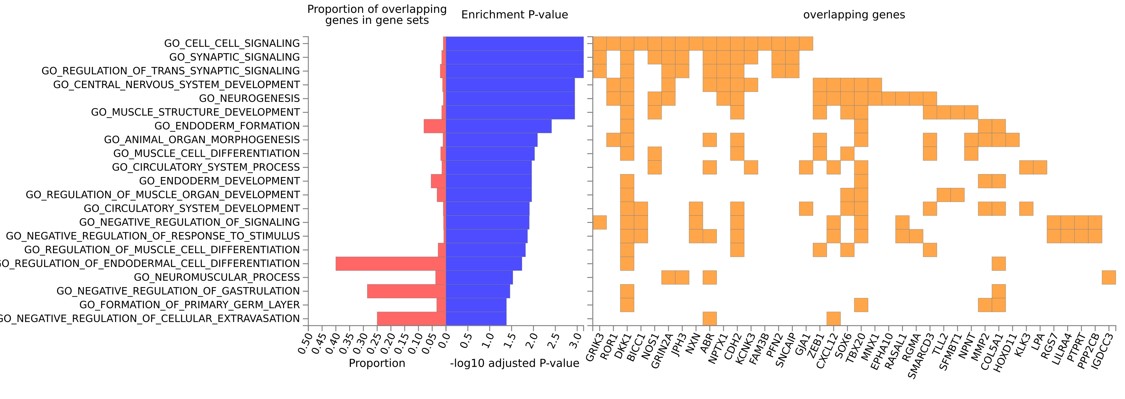

### Supplementary Figure 4

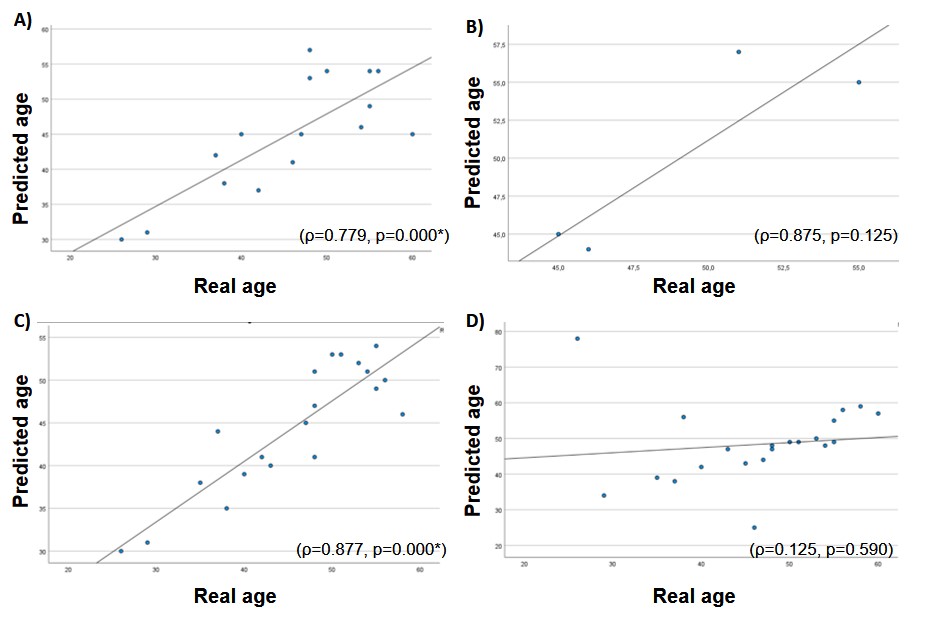

### Supplementary Text

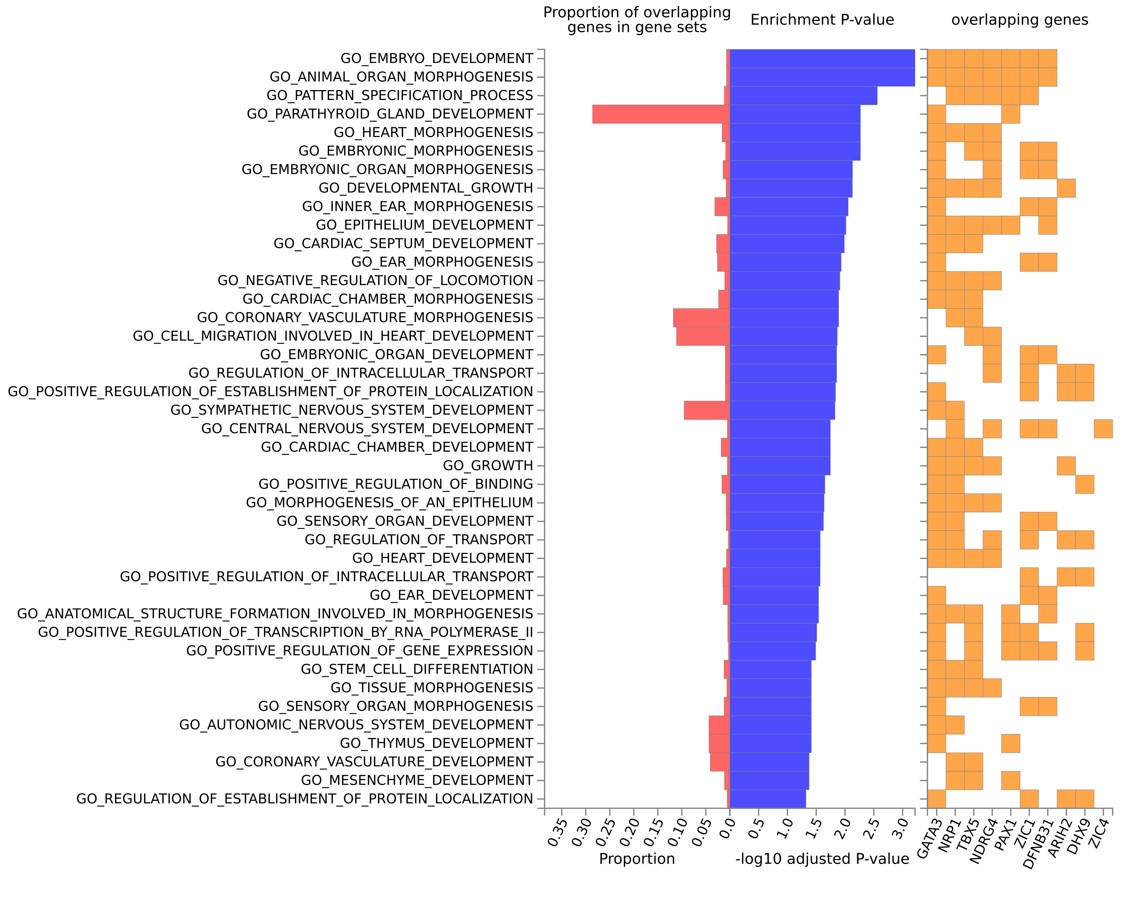
